# Relation of vaccination with severity, oxygen requirement and outcome of COVID-19 infection in Chattogram, Bangladesh

**DOI:** 10.1101/2021.06.03.21257996

**Authors:** Shuva Das, Nadia Islam Tumpa, Ayesha Ahmed Khan, Md. Minhazul Hoque, Md. Ehsanul Hoque, Safatujjahan, Kazi Farhad Ahmed, Rajat Sanker Roy Biswas

## Abstract

**Introduction:** Peoples all around the world are waiting for vaccination against COVID -19 infection. In Bangladesh, Astra-Zeneca (AZ) vaccine was provided, but patients had infections of SARS-COV-2 even after vaccination. We focused on observing the severity, oxygen requirement and outcome of the COVID-19 infected patients who took the first dose or completed the immunization regimen.

**Methods:** This is an observational study done among 174 COVID-19 patients from three COVID-19 dedicated hospitals of Chattogram, Bangladesh, who took AZ vaccines 1st dose or completed the schedule. All patients were Real-Time Reverse Transcription Polymerase Chain Reaction (rRT-PCR) positive for COVID-19. Patients were enrolled after receiving written informed consent. Suspected cases or unwilling patients were excluded from the study. Ethical approval was granted by the CMOSH–ERB. SPSS-20 was used to analyze the information gathered.

**Results:** Among 174 vaccinated patients, 55(31.61%) completed the vaccination schedule, and 119(68.39%) took their 1st dose of the COVID-19 vaccine. Gender distributions revealed 67(38.5%) female and 107(61.5%) male got the vaccine, and 55 patients completed the full two doses, and 119 patients took the 1st dose. Most of the patients were 40 years and above. In the completed vaccination group, 33(60.0%) out of 55 in and in the first dose vaccinated group, 75(63.0%) out of 119 had a mild COVID-19, and severe and critical cases were found very minimum. Among the patients who have completed the vaccination, 32(58.2%) needed no oxygen, and who was given the first dose, 78(65%) needed no oxygen. No death occurred who completed the vaccine, and 3(2.5%) patients died who took 1st dose of the vaccine.

**Conclusion:** Vaccine provided in Bangladesh to the people so far seems safe and effective. Severe and critical COVID-19 is low, and the need for oxygen to admitted patients is less, and the death rate is minimal.

## Introduction

The coronavirus disease 2019 (COVID-19) pandemic is an international public health emergency with significant social and economic disruptions and devastating health consequences. All over the world, scientists are developing vaccines. Some are already at the community level, and some are in different trial phases. The rapid development of vaccines is imperative.1

Different countries all over the world are using different vaccines from different sources. Those are showing different types of effectiveness. The immune response to many other vaccines has been shown to decrease with increasing age.2 In Bangladesh, vaccines were provided to most people who were more than 40 years early and all health care workers of different ages. Thus, the testing of SARS-CoV-2 vaccine candidates in older populations is of paramount importance since these persons account for the most serious COVID 19 cases and associated deaths.3,4 SARS-CoV-2 A study done among Health Care Workers in California5 found the rarity of positive test results 14 days after administration of the second dose of vaccine is encouraging and suggests that the efficacy of these vaccines is maintained outside the trial setting. These data underscore the critical importance of continued public health mitigation measures (masking, physical distancing, daily symptom screening, and regular testing), even in environments with a high incidence of vaccination, until herd immunity is reached at large.

## Methods

The present study is an observational study done among 174 COVID-19 patients in three COVID-19 dedicated hospitals, which are Chattogram Medical College Hospital, 250 Bedded Chattogram General Hospital and Chattagram Maa-Shishu O General Hospital from Chattogram, Bangladesh.

All patients got AZ vaccines 1st dose or completed the schedule earlier. Patients visiting the COVID-19 outdoor for testing were asked if they had received the vaccine, and if they had, they were tested for COVID-19 rRT-PCR. Again those patients who admitted to the hospital directly were asked whether they got vaccines. If the answer was yes, they were also enrolled in the study and followed up till discharge or death. All data were collected after written informed consent from the patients or the corresponding guardians, and ethical permission was obtained from the Chattagram Maa-Shishu O General Hospital’s ERB.

Patients were categorized as mild, moderate, severe and critical to define the severity according to Bangladesh’s national guideline of COVID-19 management. Also, they were divided into two groups that got oxygen in any form. Suspected cases or those who were unwilling to be included in the study were excluded. After data collection, those were analyzed by SPSS-20(IBM SPSS-Armonk, NY, USA).

## Results

Among 174 vaccinated patients, 55(31.61%) completed the full dose of COVID-19 vaccination, and 119(68.39%) took their 1^st^ dose of COVID-19 vaccine(Figure 1). Among all 67(38.5%) female and 107(61.5%) male got the vaccination, and 55 patients completed the full two-dose, and 119 patients took the 1^st^ dose(Table 1). Most of the patients are 40 years and above(Table 2) Among all 33 (60.0%) out of 55 in the completed vaccination group and 75(63.0%) out of 119 in the first dose vaccinated group were having a mild form of COVID-19 and severe and critical cases were minimum(Table 3). Who finished the immunization regimen, 32 (58.2%) of them did not require oxygen, while 78 (65%) of patients who received only the first dose did not require oxygen(Table 4) No death occurred who completed the vaccine, and 3 (2.5%) patients died who took the 1^st^ dose of vaccine(Table 5).

**Figure 1:**
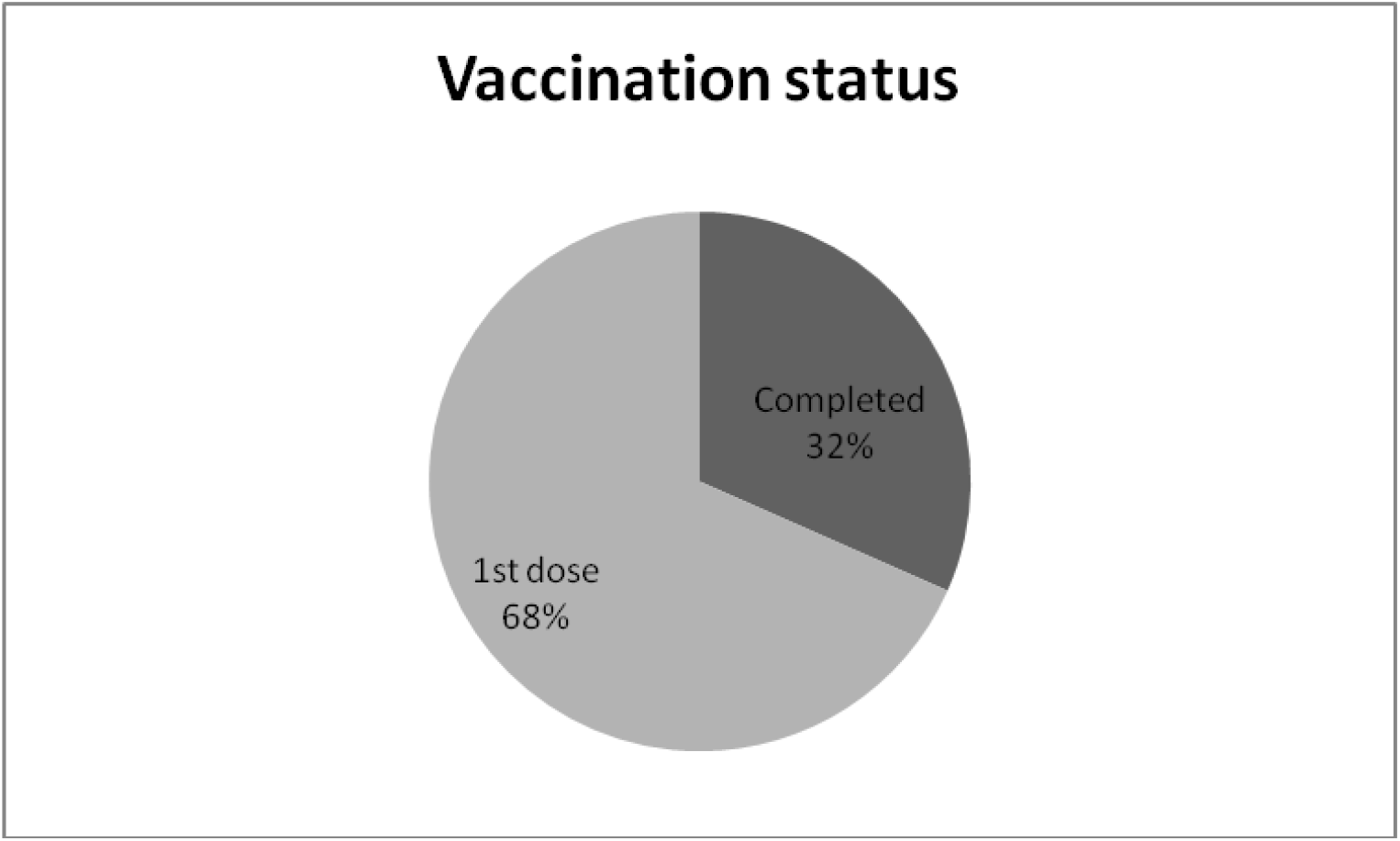
Vaccination status

**Table 1:** Vaccination and sex distribution

|  |  | Vaccination |  | Total |
| --- | --- | --- | --- | --- |
|  |  | Complete | First dose |  |
| Sex | Female | 11(20.0%) | 56(47.1%) | 67(38.5%) |
|  | Male | 44(88%) | 63(52.9%) | 107(61.5%) |
|  | Total | 55 | 119 |  |

Pearson Chi-Square value 11.63, p=0.001

**Table 2:** Vaccination and age group distribution

|  |  | Vaccination |  | Total |
| --- | --- | --- | --- | --- |
|  |  | Completed | First dose |  |
| Age<br>Group | <40 years | 8(14.5%) | 15(12.6%) | 23(13.2%) |
|  | 41-50 years | 9(16.4%) | 23(19.3%) | 32(18.4%) |
|  | 51-60 years | 13(23.6%) | 46(38.7%) | 59(33.9%) |
|  | 61-70 years | 25(45.5%) | 33(27.7%) | 58(33.3%) |
|  | >71 years | 0 | 2(1.7%) | 2(1.1%) |
| Total |  | 55 | 119 | 174 |

Pearson Chi-Square value 7.258, p=0.123

**Table 3:**
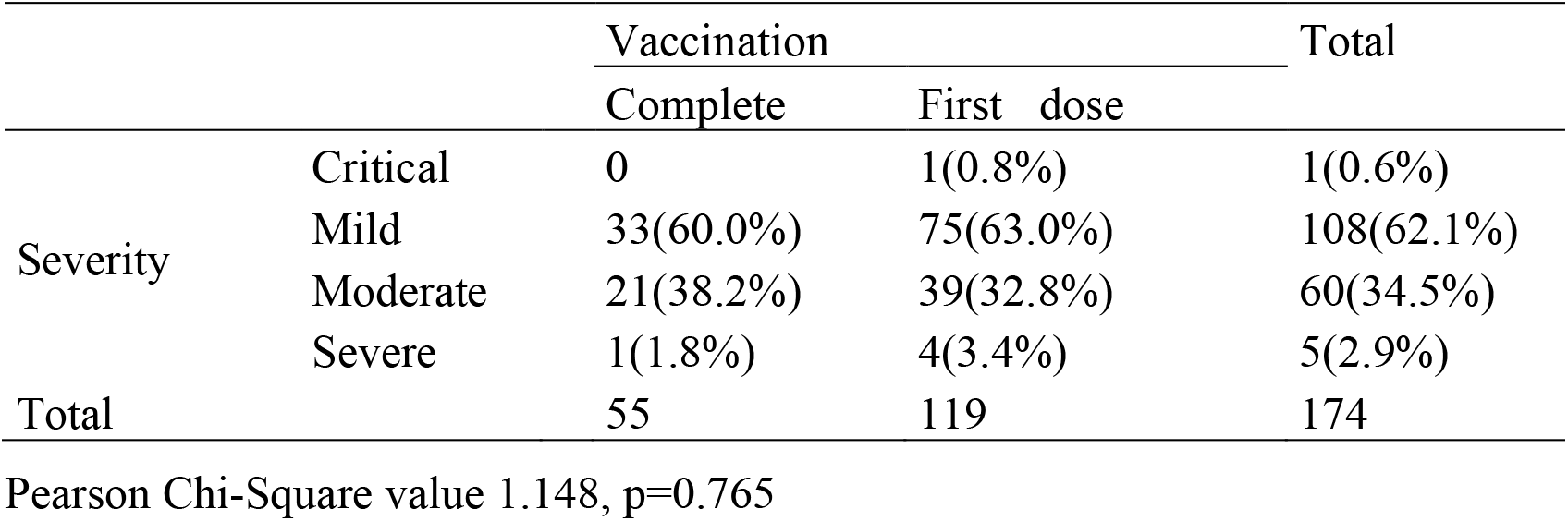
Association of Severity of COVID 19 with vaccination

|  |  | Vaccination |  | Total |
| --- | --- | --- | --- | --- |
|  |  | Complete | First dose |  |
| Severity | Critical | 0 | 1(0.8%) | 1(0.6%) |
|  | Mild | 33(60.0%) | 75(63.0%) | 108(62.1%) |
|  | Moderate | 21(38.2%) | 39(32.8%) | 60(34.5%) |
|  | Severe | 1(1.8%) | 4(3.4%) | 5(2.9%) |
| Total |  | 55 | 119 | 174 |

Pearson Chi-Square value 1.148, p=0.765

**Table 4:** Oxygen need and vaccination status

|  |  | Vaccination |  | Total |
| --- | --- | --- | --- | --- |
|  |  | Completed | First dose |  |
| Oxygen | Not needed | 32(58.2%) | 78(65.5%) | 110(63.2%) |
|  | Needed | 23(41.8%) | 41(34.5%) | 64(36.8%) |
| Total |  | 55 | 119 | 174 |

Pearson Chi-Square value 0.877, p=0.349

**Table 5:** Association of death with vaccination status

|  | Vaccination |  | Total |
| --- | --- | --- | --- |
|  | Completed | First dose |  |
| No death | 55(100%) | 116(97.5%) | 171(98.3%) |
| Death | 0 | 3(2.5%) | 3(1.7%) |
| Total | 55 | 119 | 174 |

Pearson Chi-Square value 1.411, p=0.235

## Discussion

The emergence in December 2019 of a novel coronavirus, the severe acute respiratory syndrome coronavirus 2 (SARS-CoV-2), has had devastating consequences globally.6 Control measures such as using masks, physical distancing, testing of exposed or symptomatic persons, contact tracing, and isolation have helped limit the transmission where they have been rigorously applied. However, these actions have been variably implemented and have proved insufficient in impeding the spread of COVID-19, the disease caused by SARS-CoV-2. Vaccines are needed to reduce the morbidity and mortality associated with COVID-19, and multiple vaccine platforms have been involved in the rapid development of vaccine candidates.7

In the present study, more male was vaccinated, and most of them were above 40 years. Male patients usually the earning members of the family and needs to go outside and has more chance to be infected. Again in Bangladesh, people above 40 years and health care workers of all ages got the vaccines. In an earlier study done in Bangladesh showed a similar gender distribution.8

According to our findings, merely 3 (1.7 per cent) of the 174 patients who received the full course of vaccine or only the first dose died. An outcome study has done earlier8 to see the relation of sex with a sequel where among all deaths, 40(32.2%) female died, and 323(38.08%) survived, and 84(67.8%) male died, and 525(61.92%) male survived. The total death rate is 12.75%. So vaccine has shown clearly the better effect among COVID-19 infected cases.

We found that most of the patients had mild to moderate disease, and oxygen requirement was less. But there is still a scarcity of data to compare the results in these context in different countries. A study was done in the USA9 to see the efficacy of the mRNA-1273 vaccines. It represented 94.1% efficacy at preventing Covid-19 illness, including severe disease. Aside from transient local and systemic reactions, no safety concerns were identified.

Overall, these findings show that in a small group of participants, the severity of COVID-19 was mainly mild or moderate, oxygen need among hospitalized patients was less, and the death toll was minimum provided with the AZ vaccine used in Bangladesh.

## Data Availability

Data is available in need

## Notes

### Competing Interest Statement

The authors have declared no competing interest.

### Funding Statement

No funding

### Author Declarations

IRB clearance was taken to conduct the study from the IRB of Chattogram Maa O Shishu Hospital Medical College

